# Early Prediction of Disease Progression in 2019 Novel Coronavirus Pneumonia Patients Outside Wuhan with CT and Clinical Characteristics

**DOI:** 10.1101/2020.02.19.20025296

**Authors:** Zhichao Feng, Qizhi Yu, Shanhu Yao, Lei Luo, Junhong Duan, Zhimin Yan, Min Yang, Hongpei Tan, Mengtian Ma, Ting Li, Dali Yi, Ze Mi, Hufei Zhao, Yi Jiang, Zhenhu He, Huiling Li, Wei Nie, Yin Liu, Jing Zhao, Muqing Luo, Xuanhui Liu, Pengfei Rong, Wei Wang

## Abstract

**Objective:** To determine the predictive value of CT and clinical characteristics for short-term disease progression in patients with 2019 novel coronavirus pneumonia (NCP).

**Materials and Methods:** 224 patients with confirmed 2019 novel coronavirus (COVID-19) infection outside Wuhan who had chest CT examinations were retrospectively screened. Clinical data were obtained from electronic medical records. CT images were reviewed and scored for lesion distribution, lobe and segment involvement, ground-glass opacities, consolidation, and interstitial thickening. All included patients with moderate NCP were observed for at least 14 days from admission to determine whether they exacerbated to severe NCP (progressive group) or not (stable group). CT and clinical characteristics between the two groups were compared, and multivariate logistic regression and sensitivity analyses were performed to identify the risk factors for developing severe NCP.

**Results:** A total of 141 patients with moderate NCP were included, of which 15 (10.6%) patients developed severe NCP during hospitalization and assigned to the progressive group. Multivariate logistic regression analysis showed that higher neutrophil-to-lymphocyte ratio (NLR) (odds ratio [OR] and 95% confidence interval [CI], 1.26 [1.04-1.53]; *P* = 0.018) and CT severity score (OR and 95% CI, 1.25 [1.08-1.46]; *P* = 0.004) on admission were independent predictors for progression to severe NCP, and sensitivity analysis confirmed the consistent results in nonimported patients but not in imported patients. However, no significant difference in lung involvement was found on CT between imported and nonimported patients (all *P* > 0.05). Patients who were admitted more than 4 days from symptom onset tended to have more severe lung involvement. Spearman correlation analysis showed the close association between CT severity score and inflammatory indexes (*r* = 0.17∼0.47, all *P* < 0.05).

**Conclusion:** CT severity score was associated with inflammatory levels and higher NLR and CT severity score on admission were independent risk factors for short-term progression in patients with NCP outside Wuhan. Furthermore, early admission and surveillance by CT should be recommended to improve clinical outcomes.

## Introduction

The outbreak of 2019 novel coronavirus pneumonia (NCP) originated from Wuhan has shown the ability of human-to-human transmission and rapidly spread to become a world-wide emergency along with increasing imported and secondary contacted infection risk.^1^ Most patients with NCP have a mild clinical course, while a proportion of patients demonstrated rapid deterioration (particularly within 7-14 days) from onset of symptoms into severe NCP with or without acute respiratory distress syndrome (ARDS). These patients have poor survival and often require intensive medical resource utilization, and the mortality of them are about 20 times higher than that of non-severe 2019 novel coronavirus (COVID-19) patients. ^2,3^ Thus, early identification of patients at risk of serious complications of NCP is of clinical importance. Several studies reported that the prevalence of severe NCP ranged from 15.7% to 26.1% and these cases were often associated with abnormal chest CT findings and clinical laboratory data.^3-5^ Guan et al indicated that patients with severe NCP were more likely to show ground-glass opacity (GGO), local or bilateral patchy shadowing, and interstitial abnormalities on CT.^5^ This likely reflects the clinical progression of disease but also offers an opportunity to investigate the clinical utility of chest CT as a predictive tool to risk-stratify the patients. Furthermore, the predictive value of chest CT in NCP prognosis is warranted as to assist the effective treatment and control of disease spread. Previous study suggested that higher CT lung score correlated with poor prognosis in patients with Middle East respiratory syndrome coronavirus (MERS).^6^ Chest CT has been demonstrated to be an important approach for screening individuals with suspected NCP and monitoring treatment response according to the dynamic radiological changes of NCP.^7^ Therefore, we enrolled a cohort of patients with moderate NCP on admission and observed for at least 14 days to explore the early CT and clinical risk factors for progression to severe NCP. Meanwhile, we also compared the CT and inflammatory indexes in patients with different source of infection or period from symptom onset to admission to provide deep understanding of the relationship among CT findings, epidemiological features, and inflammation.

## Materials and Methods

### Patients

Our study was conducted in compliance with the Health Insurance Portability and Accountability Act (HIPAA). The permission of Institutional Review Board of The Third Xiangya Hospital was obtained for retrospective assessment of clinical and radiological data with waiver of informed consent.

Health records were reviewed for patients who were treated at Third Xiangya Hospital, Changsha Public Health Treatment Center, and Second People’ s Hospital of Hunan between January 17, 2020 and February 1, 2020. Patients were included in the study if they satisfied the following criteria: 1) confirmed COVID-19 infection; 2) available chest CT scan on admission. The diagnosis of COVID-19 infection was established based on the World Health Organization (WHO) interim guidance, and a confirmed case was defined as a positive result to high-throughput sequencing or real-time reverse transcription-polymerase chain reaction (RT-PCR) assay for nasal and pharyngeal swab specimens. Patients were observed for at least 14 days from admission to determine whether they exacerbated to severe NCP (progressive group) or not (stable group).

### Clinical data collection

Demographic, epidemiological, and laboratory data were obtained with data collection forms from electronic medical records. According to the source of infection, patients were divided into imported group (patients who had recently been to Wuhan) and nonimported group (patients who contacted with people from Wuhan or local infected patients). The clinical classification of NCP are as follows: (1) moderate type, patients with fever, respiratory tract symptoms, and radiological evidence of confirmed pneumonia. (2) severe type, patients with one of the following: a) respiratory distress (respiratory rate ≥ 30 beats/min); b) hypoxia (oxygen saturation ≤ 93% in the resting state); c) hypoxemia (arterial blood oxygen partial pressure/oxygen concentration ≤ 300mmHg). (3) critical type, patients with one of the following: a) respiratory failure requiring mechanical ventilation; b) shock; c) intensive care unit (ICU) admission is required for combined other organs failure. In this study, severe NCP broadly included severe and critical types as above defined. The endpoint of this study was the development of severe NCP, and patients who discharged or did not developed severe NCP at enrollment were censored.

### CT examination and image analysis

The SOMATOM go.Top scanner (Siemens, Erlangen, Germany), GE Revolution CT scanner (GE Healthcare, Waukesha, USA), and TOSHIBA Aquilion 16 (Toshiba, Tokyo, Japan) were used for chest CT examinations. The images were reconstructed to 1.0-mm thickness for the transverse scans. Sagittal and coronal reconstructions with a 3.0-mm thickness were performed.

All CT images were reviewed independently by two radiologists, each with over 10 years of experience in chest imaging. A third experienced radiologist was consulted if there was a disagreement in interpreting imaging results. The imaging features including ground-glass opacities (GGO), consolidation, crazy-paving, and air bronchogram defined as previous studies were recorded.^8,9^ The lesion distribution pattern, lobe and segment involvement were also assessed. The CT findings in the outer one third of the lung were defined as peripheral, and those in the inner two thirds of the lung were defined as central. Besides, the presence of discrete nodules, lymphadenopathy, and pleural effusion was recorded. Each of the five lung lobes was reviewed for opacification and consolidation. The lesions extent within each lung lobe was semi-quantitatively evaluated by scoring from 0 to 5 based on the degree of involvement: score 0, none involvement; score 1, ≤ 5% involvement; score 2, 6%∼25% involvement; score 3, 26%∼50% involvement; score 4, 51%∼75% involvement; score 5, > 75% involvement. The total score was calculated by summing up scores of all five lobes to provide a CT severity score ranging from 0 to 25.^7^

### Statistical Analysis

Quantitative variables are presented as median and interquartile range (IQR), and categorical variables are presented as frequency and percentage. Normality of the continuous variables was assessed with the Kolmogorov–Smirnov test. Differences between groups were examined using Student’ s t-test or Mann-Whitney U test for quantitative variables according to the normal distribution and Chi-square test or Fisher’ s exact test for categorical variables. Multivariate logistic regression with forward stepwise selection based on likelihood ratio was used to identify the risk factors for development of severe NCP, and sensitivity analysis was performed according to the source of infection. Statistical analyses were performed using IBM SPSS statistics software (version 22.0, SPSS Inc., Chicago, IL, USA) and R software (version 3.6.1). A *P* value of less than 0.05 considered to be statistically significant.

## Results

### 1. Clinical characteristics

A total of 224 patients were identified according to the inclusion criteria, of which 83 patients were excluded for having: 1) observation period from admission less than 14 days (n = 60); 2) negative CT findings or severe NCP on admission (n = 16); 3) age younger than 18 years old (n = 7). Finally, a cohort of 141 patients were included in our study (Figure 1). The baseline demographic, epidemiological, and laboratory characteristics of included 141 patients are presented in Table 1. Among them, 76 (53.9%) patients who had recently been to Wuhan were imported and 72 (51.1%) were male, with a median age of 44 years and a median period symptom onset to admission of 4 days. 52 patients of them had discharged at enrollment.

**Table 1.** Baseline clinical and CT characteristics of patients with NCP

|  | All (n = 141) | Stable (n = 126) | Progressive (n= 15) | P value |
| --- | --- | --- | --- | --- |
| Age (years) | 44 (34-55) | 41 (33-52) | 58 (44-66) | **0.001** |
| Male gender | 72 (51.1) | 65 (51.6) | 7 (46.7) | 0.719 |
| Exposure within 14 days |  |  |  | 0.616 |
| Imported | 76 (53.9) | 67 (53.2) | 9 (60.0) |  |
| Nonimported | 65 (46.1) | 59 (46.8) | 6 (40.0) |  |
| Period from symptom onset to admission (days) | 4 (2-7) | 5 (2-7) | 4 (2-5) | 0.200 |
| Smoking history | 7 (5.0) | 7 (5.6) | 0 (0) | 0.349 |
| Comorbidity |  |  |  |  |
| Any | 33 (23.4) | 26 (20.6) | 7 (46.7) | **0.024** |
| Diabetes | 8 (5.7) | 6 (4.8) | 2 (13.3) | 0.175 |
| Hypertension | 21 (14.9) | 15 (11.9) | 6 (40.0) | **0.004** |
| Cardiovascular disease | 3 (2.1) | 2 (1.6) | 1 (6.7) | 0.288 |
| COPD | 4 (2.8) | 2 (1.6) | 2 (13.3) | 0.056 |
| Cerebrovascular disease | 1 (0.7) | 1 (0.8) | 0 (0) | 0.729 |
| Hepatitis B infection | 4 (2.8) | 4 (3.2) | 0 (0) | 0.484 |
| Signs and symptoms |  |  |  |  |
| Fever | 105 (74.5) | 92 (73.0) | 13 (86.7) | 0.252 |
| Cough | 74 (52.5) | 66 (52.4) | 8 (53.3) | 0.944 |
| Sputum production | 16 (11.3) | 14 (11.1) | 2 (13.3) | 0.798 |
| Fatigue | 29 (20.6) | 26 (20.6) | 3 (20.0) | 0.954 |
| Headache | 5 (3.5) | 4 (3.2) | 1 (6.7) | 0.435 |
| Anorexia | 5 (3.5) | 3 (2.4) | 2 (13.3) | 0.088 |
| Diarrhea | 6 (4.3) | 4 (3.2) | 2 (13.3) | 0.065 |
| Laboratory findings |  |  |  |  |
| Platelet count ($\times 10^9/L$) | 188.5 (134.8-240.0) | 190.5 (135.8-240.0) | 152.5 (114.5-220.3) | 0.383 |
| White blood cell count ($\times 10^9/L$) | 4.6 (3.4-6.3) | 4.5 (3.4-6.3) | 4.6 (3.3-7.0) | 0.843 |
| Neutrophil count ($\times 10^9/L$) | 3.4 (2.3-4.3) | 3.4 (2.2-4.3) | 3.2 (2.6-5.2) | 0.300 |
| Lymphocyte count ($\times 10^9/L$) | 1.1 (0.7-1.6) | 1.2 (0.8-1.6) | 0.7 (0.6-1.0) | **0.008** |
| NLR | 3.4 (2.1-4.2) | 3.4 (1.9-3.9) | 5.2 (3.1-6.9) | **0.001** |
| Albumin (g/L) | 38.0 (32.7-39.2) | 38.1 (32.9-39.5) | 33.4 (30.6-36.0) | 0.084 |
| Creatinine ($\mu\text{mol/L}$) | 73.3 (65.4-75.5) | 73.3 (63.9-75.5) | 70.0 (67.0-97.7) | 0.581 |
| C-reactive protein (mg/L) | 21.7 (9.4-33.5) | 19.2 (8.9-24.8) | 44.0 (25.0-51.6) | **0.002** |
| CT features |  |  |  |  |
| Number of lobes involved |  |  |  | **0.013** |
| One lobe | 14 (10.0) | 12 (9.5) | 2 (13.3) |  |
| Two lobes | 25 (17.7) | 25 (19.9) | 0 (0) |  |
| Three lobes | 15 (10.6) | 15 (11.9) | 0 (0) |  |
| Four lobes | 29 (20.6) | 28 (22.2) | 1 (6.7) |  |
| Five lobes | 58 (41.1) | 46 (36.5) | 12 (80.0) |  |
| Number of segments involved | 9 (5-12) | 9 (5-12) | 14 (12-15) | **0.001** |
| Bilateral involvement | 123 (87.2) | 110 (87.3) | 13 (86.7) | 0.944 |
| Distribution pattern |  |  |  | 0.116 |
| Peripheral | 71 (50.4) | 67 (53.2) | 4 (26.7) |  |
| Central | 2 (1.4) | 2 (1.6) | 0 (0) |  |
| Mixed | 68 (48.2) | 57 (45.2) | 11 (73.3) |  |
| GGO | 135 (95.7) | 120 (88.9) | 15 (100) | 0.388 |
| Consolidation | 120 (85.1) | 107 (84.9) | 13 (86.7) | 0.858 |
| GGO with consolidation | 113 (80.1) | 100 (79.4) | 13 (86.7) | 0.503 |
| Crazy-paving | 42 (29.8) | 34 (27.0) | 8 (53.3) | **0.035** |
| Air bronchogram | 82 (58.2) | 71 (56.4) | 11 (73.3) | 0.207 |
| Discrete nodules | 11 (7.8) | 10 (7.9) | 1 (6.7) | 0.862 |
| Lymphadenopathy | 6 (4.3) | 5 (4.0) | 1 (6.7) | 0.625 |
| Pleural effusion | 4 (2.8) | 4 (3.2) | 0 (0) | 0.484 |
| CT severity score | 6 (4-10) | 6 (3-9) | 10 (7-15) | **0.003** |

COPD, chronic obstructive pulmonary disease; CT, computed tomography; GGO, ground-glass opacities; NCP, 2019 novel coronavirus pneumonia; NLR, neutrophil-to-lymphocyte ratio.

**Figure 1.**
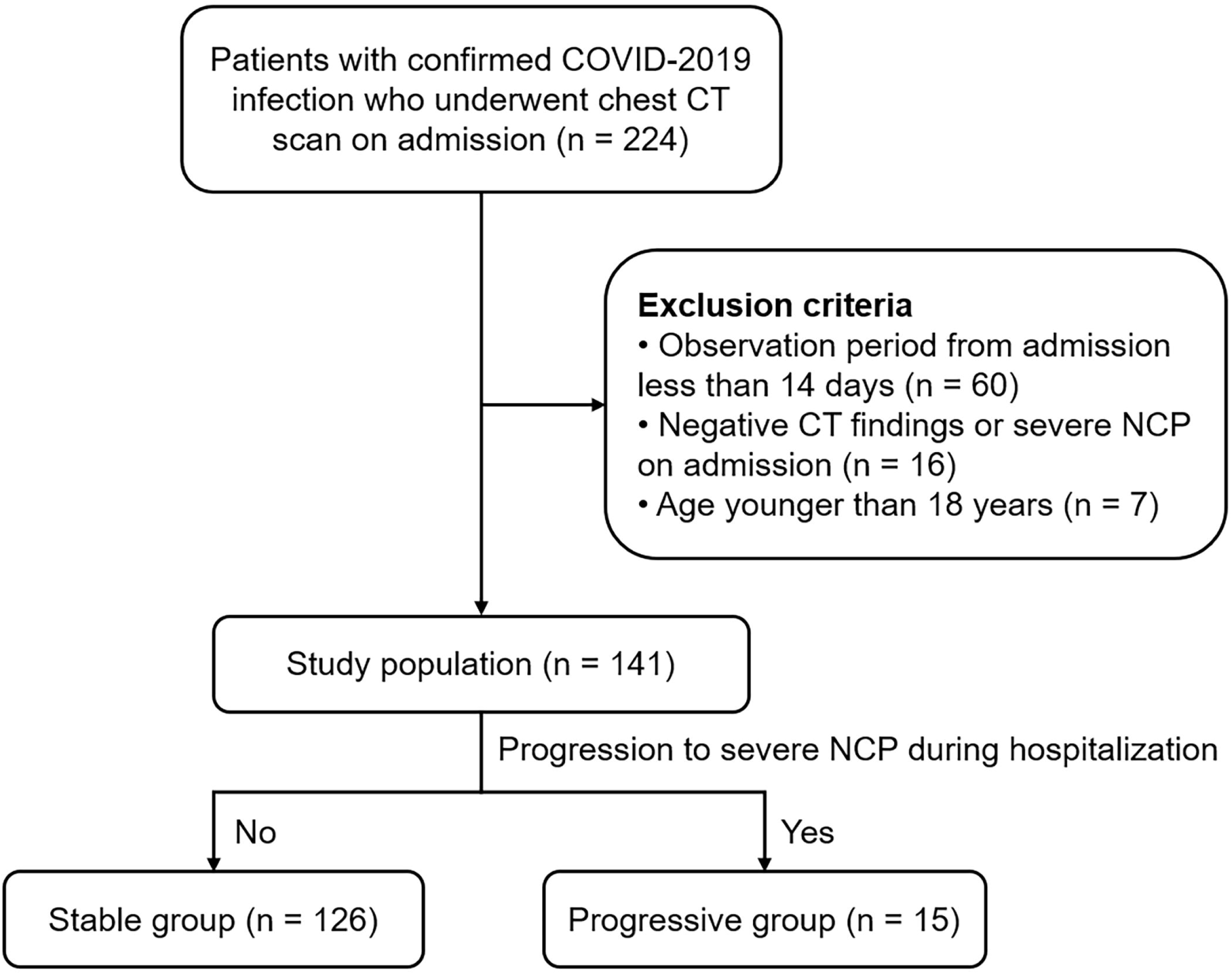
Flow diagram shows the study population enrollment and observation period.

During the hospitalization, 15 cases progressed to severe NCP (progressive group) and the remaining patients did not (stable group). Compare with stable group, patients in progressive group were significantly older (*P* = 0.001), but the male gender and imported proportions were not shown statistical difference. Patients who progressed to severe NCP were more likely to have underlying hypertension (*P* = 0.004), but did not otherwise have significant differences in other co-morbidities including diabetes, chronic obstructive pulmonary disease (COPD), cardiovascular disease, cerebrovascular disease and chronic hepatitis B infection. The main symptoms between the two groups were not statistically different, while slightly more patients manifested digestive symptoms in progressive group, such as anorexia and diarrhea (*P* = 0.088 and 0.065, respectively). Patients in the progressive group had lower baseline lymphocyte count, higher neutrophil-to-lymphocyte ratio (NLR), and C-reactive protein (all *P* < 0.01).

### 2. CT findings

There were 87 (61.7%) patients with 4 to 5 lobes involved, with a median of 9 segments involved. Most lesions were bilateral (87.2%) and peripheral or mixed distributed (98.6%). The main CT characteristics of patients with NCP included GGO, GGO with crazy-paving, consolidation, and GGO with consolidation, ranging from 29.8%∼95.7% (Table 1; Figure 2 a-f). Other infrequent features included discrete nodules, lymphadenopathy, and small pleural effusion. Compared with stable group, patients in progressive group had more lobes and segments involved, higher proportion of crazy-paving sign, and higher CT severity score (all *P* < 0.05; Figure 3 a).

**Figure 2.**
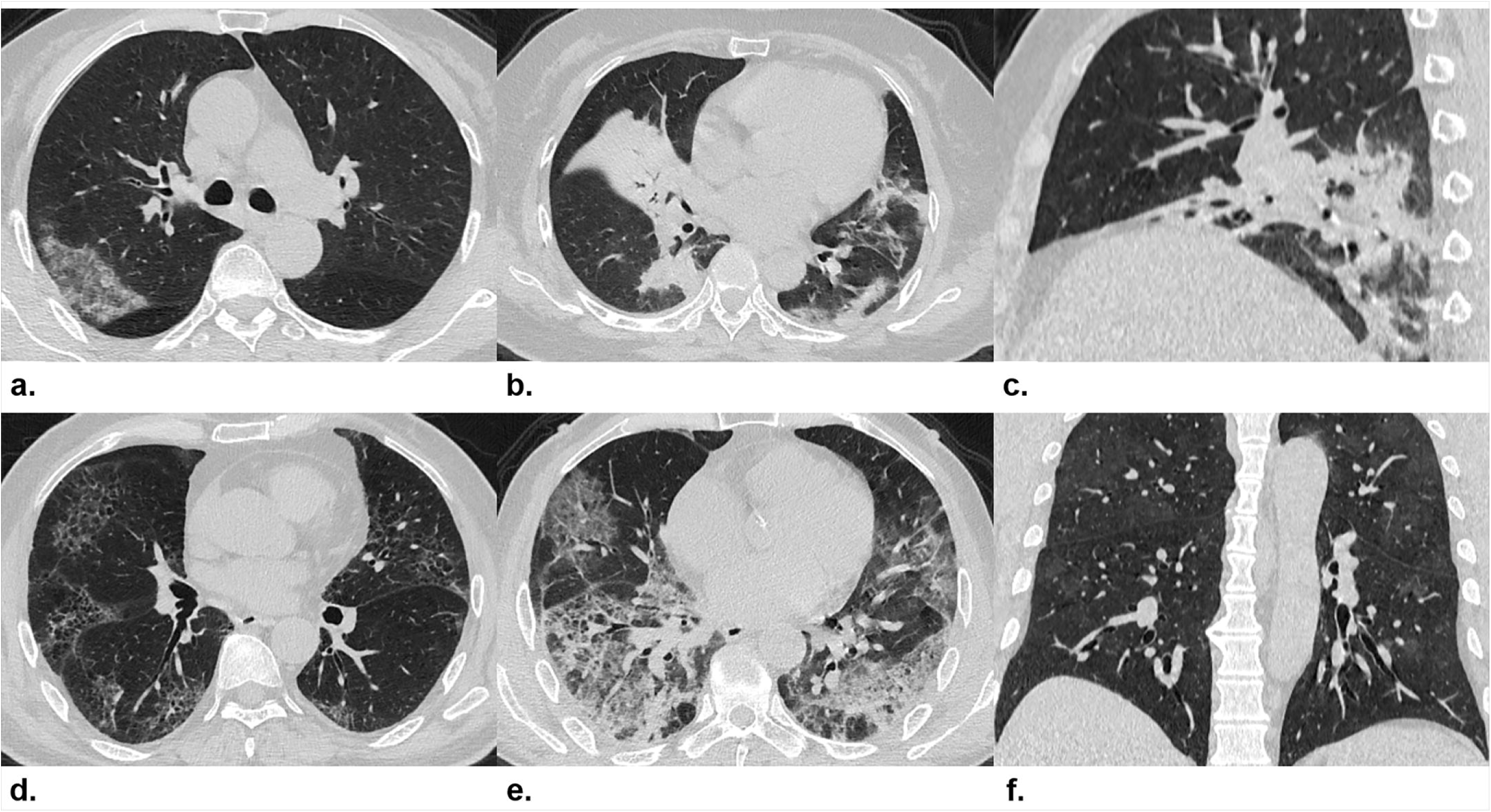
(a) Subpleural patchy areas of GGO with crazy-paving sign in right middle lobe. (b) Multiple patchy areas of consolidation in right middle lobe, left upper lobe, and bilateral lower lobes and air bronchogram in right middle lobe. (c) Multiple patchy areas of organizing pneumonia in right middle and lower lobes on sagittal image with CT severity score of 9 for right lung. (d) Bilateral and peripheral multiple patchy areas of GGO with reticular and intralobular septal thickening. (e) Multiple mixed distributed pure GGO, GGO with consolidation, and interlobular septal thickening in bilateral lungs. (f) Bilateral multiple patchy and thin areas of GGO in posterior parts of the lungs. GGO, ground-glass opacities.

**Figure 3.**
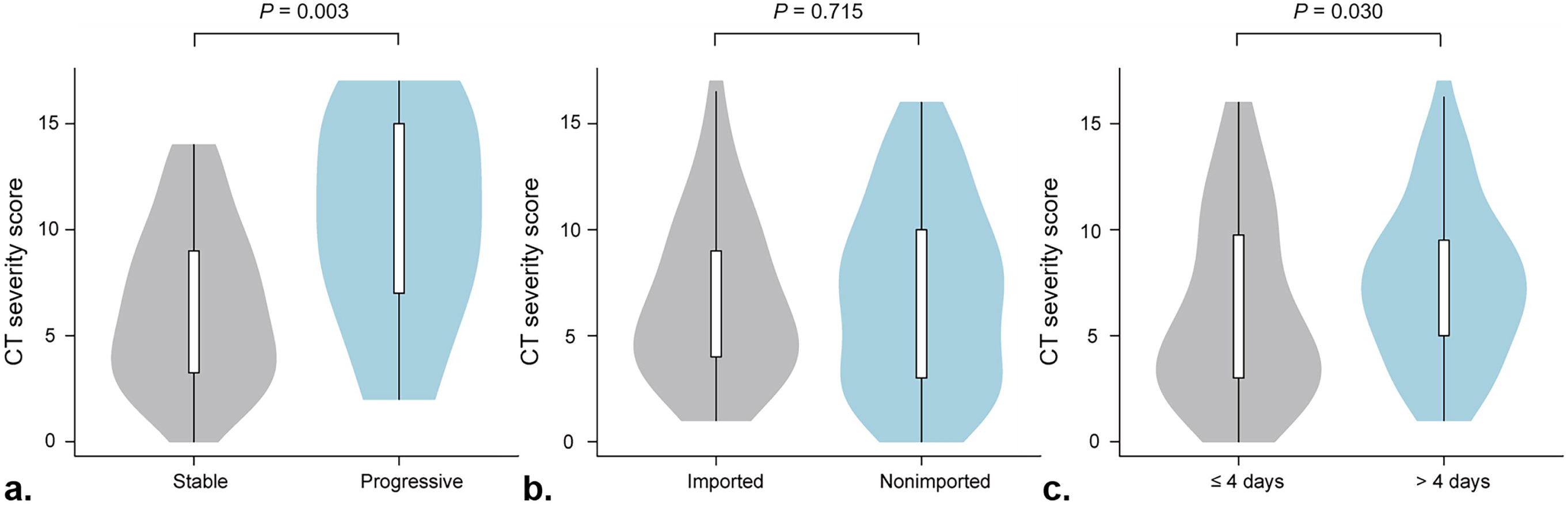
The boxplots of the CT severity score between stable group and progressive group (a), between imported patients and nonimported patients (b), and between the patients who were admitted ≤ 4 days and > 4 days from symptom onset.

### 3. Multivariate logistic regression and sensitivity analyses

Logistic regression analysis showed that baseline NLR (odds ratio [OR] and 95% confidence interval [CI], 1.26 [1.04-1.53]; *P* = 0.018) and CT severity score (OR and 95% CI, 1.25 [1.08-1.46]; *P* = 0.004) were independent predictors for progression to severe NCP (Table 2). When stratifying by the source of infection (imported or nonimported), the results showed that baseline NLR (OR and 95% CI, 1.71 [1.05-2.79]; *P* = 0.023) and CT severity score (OR with 95% CI, 1.58 [1.09-2.28]; *P* = 0.001) remained the significant risk factors in nonimported patients, but not in imported patients. Among the imported patients, only age (OR with 95% CI, 1.13 [1.04-1.22]; *P* = 0.004) was the predictor for progression to severe NCP.

**Table 2.**
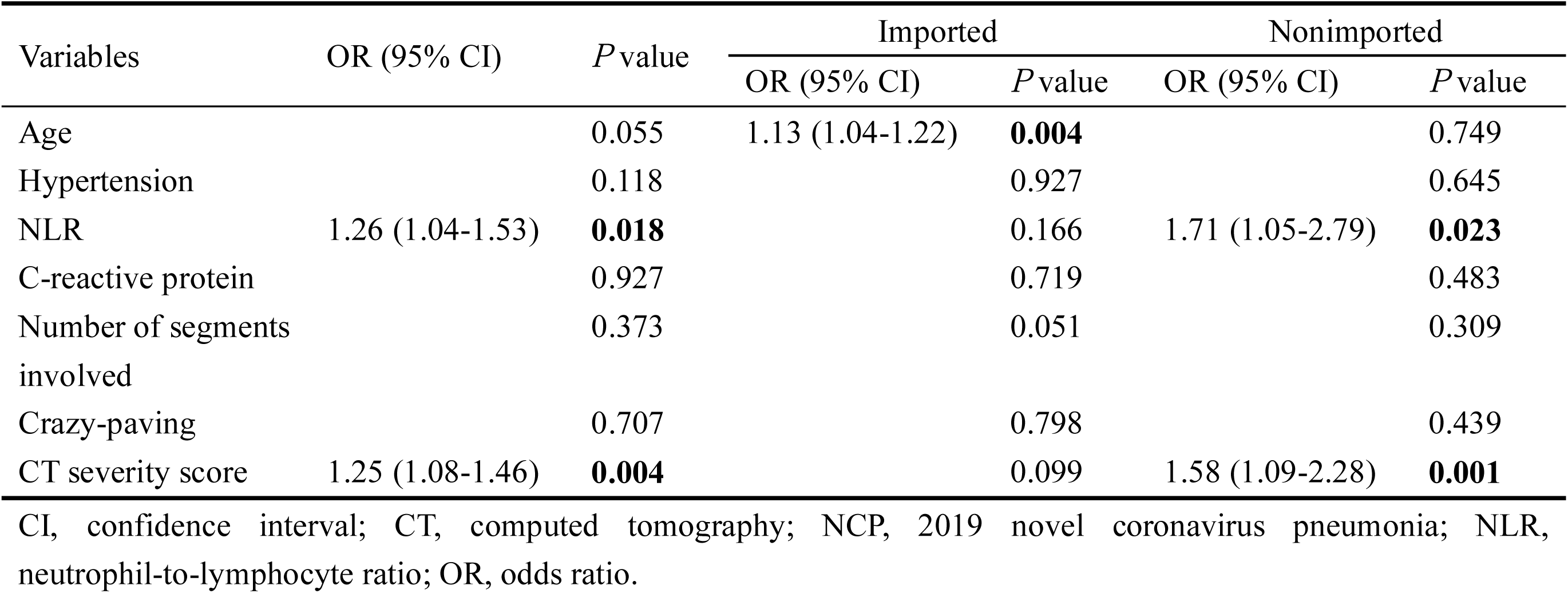
Risk factors for progression to severe NCP by multivariate logistic regression and sensitive analyses

| Variables | OR (95% CI) | *P* value | Imported |  | Nonimported |  |
| --- | --- | --- | --- | --- | --- | --- |
|  |  |  | OR (95% CI) | *P* value | OR (95% CI) | *P* value |
| Age |  | 0.055 | 1.13 (1.04-1.22) | **0.004** |  | 0.749 |
| Hypertension |  | 0.118 |  | 0.927 |  | 0.645 |
| NLR | 1.26 (1.04-1.53) | **0.018** |  | 0.166 | 1.71 (1.05-2.79) | **0.023** |
| C-reactive protein |  | 0.927 |  | 0.719 |  | 0.483 |
| Number of segments involved |  | 0.373 |  | 0.051 |  | 0.309 |
| Crazy-paving |  | 0.707 |  | 0.798 |  | 0.439 |
| CT severity score | 1.25 (1.08-1.46) | **0.004** |  | 0.099 | 1.58 (1.09-2.28) | **0.001** |

CI, confidence interval; CT, computed tomography; NCP, 2019 novel coronavirus pneumonia; NLR, neutrophil-to-lymphocyte ratio; OR, odds ratio.

**Table 3.** Comparison of CT characteristics between imported and nonimported patients with NCP

|  | Imported (n=76) | Nonimported (n=65) | *P* value |
| --- | --- | --- | --- |
| Number of lobes involved | 4 (2-5) | 4 (2-5) | 0.799 |
| Number of segments involved | 9 (4-12) | 9 (5-13) | 0.383 |
| Bilateral involvement | 66 (86.8) | 57 (87.7) | 0.880 |
| Distribution pattern |  |  | 0.988 |
| Peripheral | 38 (50.0) | 33 (50.8) |  |
| Central | 1 (1.3) | 1 (1.5) |  |
| Mixed | 37 (48.7) | 31 (47.7) |  |
| GGO | 74 (97.4) | 61 (93.9) | 0.302 |
| Consolidation | 66 (86.8) | 54 (83.1) | 0.531 |
| GGO with consolidation | 63 (82.9) | 50 (76.9) | 0.376 |
| Crazy-paving | 22 (29.0) | 20 (30.8) | 0.814 |
| Air bronchogram | 46 (60.5) | 36 (55.4) | 0.537 |
| Discrete nodules | 4 (5.3) | 7 (10.8) | 0.224 |
| Lymphadenopathy | 3 (4.0) | 3 (4.6) | 1.000 |
| Pleural effusion | 1 (1.3) | 3 (4.6) | 0.335 |
| CT severity score | 6 (4-9) | 7 (3-10) | 0.715 |

CT, computed tomography; GGO, ground-glass opacities; NCP, 2019 novel coronavirus pneumonia.

**Table 4.** Comparison of clinical and CT characteristics according to the period from symptom onset to admission

|  | ≤ 4 days (n=70) | > 4 days (n=71) | *P* value |
| --- | --- | --- | --- |
| Age (years) | 41 (31-53) | 46 (35-59) | 0.147 |
| White blood cell count (×10<sup>9</sup>/L) | 5.0 (3.6-6.4) | 4.2 (3.3-6.2) | 0.277 |
| NLR | 3.4 (2.6-4.9) | 3.4 (1.8-3.9) | 0.074 |
| C-reactive protein (mg/L) | 23.4 (10.4-39.4) | 18.8 (8.0-28.4) | 0.215 |
| Number of lobes involved | 3 (2-5) | 4 (3-5) | **0.010** |
| Number of segments involved | 7 (3-12) | 10 (6-12) | **0.020** |
| Crazy-paving | 25 (35.7) | 17 (23.9) | 0.126 |
| CT severity score | 5 (3-10) | 7 (5-10) | **0.030** |

CT, computed tomography; NLR, neutrophil-to-lymphocyte ratio.

### 4. Comparison between subgroups according to source of infection and period from symptom onset to admission

There was no significant difference in the CT findings between imported and nonimported patients, who had similar lesion distribution pattern, main imaging features and comparable lung involvement and CT severity score (Figure 3 b). For patients who were admitted more than 4 days from symptom onset, more lobes or segments involved and higher CT severity score were found (Figure 3 c), while there was no significant difference in age and inflammatory indexes (including white blood cell count, NLR, and C-reactive protein) between patients with different period (≤ 4 days vs. > 4 days) from symptom onset to admission.

Further Spearman correlation analysis showed the close association between CT characteristics and inflammatory indexes, especially for CT severity score and C-reactive protein (Figure 4).

**Figure 4.**
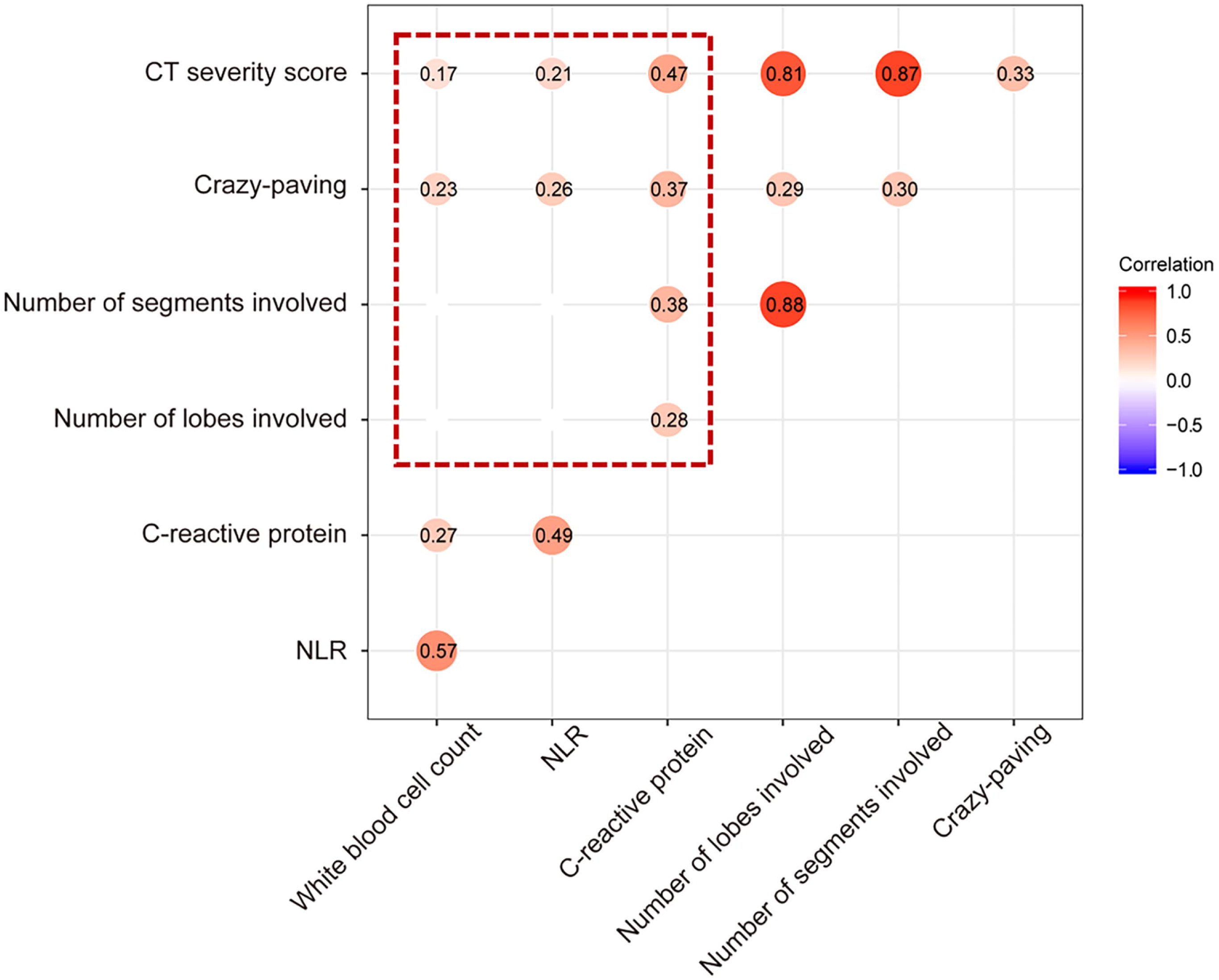
Heatmap depicting the correlations between inflammatory markers and CT characteristics (within the red dotted box). It shows the correlation coefficients *r* with *P* < 0.05 of all pairs. It can be observed that C-reactive protein and CT severity score had the strongest correlation. CT, computed tomography.

## Discussion

In this study, we retrospectively assessed the CT and clinical characteristics of 141 patients with NCP outside Wuhan and identified the baseline risk factors for clinical progression. Our results showed that higher NLR and CT severity score on admission were independent predictors for progression to severe NCP, especially in nonimported patients. In addition, patients who were admitted longer from symptom onset tended to have more severe lung involvement.

With the increase of newly confirmed and severe cases, the management of severe patients brings huge challenge in efficiently controlling the COVID-19 outbreak. Some patients progressed rapidly with ARDS and subsequent multiple organ dysfunction syndrome (MODS).^2^ Early identification of patients with high risk to develop ARDS or MODS and risk stratification management might be helpful to reduce mortality. In our cohort, the prevalence of severe NCP was 10.6%, which was lower than some large-scale reports.^3,5^ The main cause was because our patients were moderate on admission and enrolled from one single city. We found that progressive patients were older and had a greater number of underlying hypertension than stable patients. These data were in agreement with another recent report, which suggested that age and history of hypertension may be risk factors for short-term progression.^10^ Our study further showed that progressive patients had lower baseline lymphocyte count, higher NLR and C-reactive protein. COVID-19 virus might act on lymphocytes as does severe acute respiratory syndrome coronavirus (SARS-CoV) which induces a cytokine storm and results in a series of immune responses.^11^ Some studies suggest that decrease of T lymphocyte count indicate that coronavirus consumes many immune cells and inhibits the cellular immune function, and reduced but hyperactivated peripheral T cells partially accounts for the severe immune injury in COVID-19 patients.^12,13^ Thus, damage to lymphocytes might be an important factor leading to exacerbations of patients. NLR, as a simple lymphocyte related parameter to assess easily the inflammatory status, is widely used for the prediction of prognosis of patients with pneumonia.^14,15^ Higher NLR indicates damaged lymphocyte function and/or risk of bacterial infection due to low immune function. In addition, C-reactive protein is another serum maker produced by the liver in response to inflammation. Liu et al showed that C-reactive protein might be predictive of disease severity in COVID-19 infected patients. ^16^ Thus, our results suggested that patients with higher inflammatory levels on admission had higher risk to develop severe NCP.

To explore the predictive value of CT for progression, we compared the difference of CT characteristics in stable and progressive patients, and found that progressive patients had greater lung involvement and higher CT severity score. CT severity score is used to semi-quantitatively estimate the pulmonary involvement, which is associated with both number of involved lobes and extent of lesions. ^17^ Pan et al reported a 4-stage dynamic pattern of NCP based on CT severity score and indicated that lung abnormalities showed greatest severity approximately 10 days after initial onset of symptoms. ^7^ In support of our findings, a previous report regarding MERS showed the predictive value of CT severity score for prognosis and short-term mortality.^6^ Furthermore, higher proportion of progressive patients showed crazy-paving sign which reflects interstitial thickening. ^18^ Guan et al also reported higher proportion of severe patients with interstitial abnormalities.^5^ The binding of COVID-19 spike protein to the receptor angiotensin converting enzyme II (ACE2) contributes to the downregulation of ACE2, increased pulmonary capillary permeability, and diffuse alveolar damage during NCP.^19,20^ In patients with SARS, mixed and predominant reticular patterns were noted from the second week.^8,21^ Thus, we speculated that the involvement of the interstitial vascular endothelial cells results in interlobular and intralobular septal thickening, which is associated with the disease severity.

Our results showed that NLR and CT severity score on admission were significant predictors for progression to severe illness. The early predictive value of NLR has been reported in a recent study.^10^ To our knowledge, this is the first study to indicate the role of CT to predict disease progression in patients with NCP. Previous study showed that the MuLBSTA score can early warn the mortality of viral pneumonia, which included lymphopenia and multilobe infiltration.^22^ Our findings were consistent with theirs but more quantitatively in terms of imaging evaluation of lung involvement. Like SARS and MERS, some NCP patients progressed rapidly at about 10-14 days after onset likely dues to the cytokine storm in the body as evidenced by increased plasma proinflammatory cytokines.^11,23^ Our results further revealed the significant association between inflammatory markers and CT characteristics, especially for C-reactive protein and CT severity score, which indicated the potential value of CT to estimate pulmonary inflammation and lung damage. Furthermore, the predictive significance of NLR and CT severity score was confirmed only in nonimported patients outside Wuhan. Among the imported patients from Wuhan, the elder had higher risk to develop severe illness. However, the reason for this discrepancy between imported and nonimported patients remains to be elucidated.

The patients who were admitted more than 4 days after symptom onset had more severe lung involvement, which likely attributed to the lung involvement progression as disease course extends. Song et al found that more consolidation and less GGO lesions in patients with an interval > 4 days between symptom onset and CT scan.^24^ Thus, early admission and surveillance by CT is crucial for the individual management of patients with NCP.

There were some limitations in our study. First, our study retrospectively included 141 patients with moderate NCP on admission from one single city outside Wuhan. Data from multiple cities are needed to further validate our findings. Second, dynamic inflammatory indexes and treatment during hospitalization were not included in the analysis. More comprehensive investigation of the relationship between CT characteristics and cytokine storm induced by COVID-2019 needs to be performed.

In conclusion, our results showed that higher NLR and CT severity score on admission were independent risk factors for short-term progression in patients with NCP. The CT findings in patients with NCP were associated with inflammatory levels and both them have the potential to predict disease progression. Imported and nonimported patients outside Wuhan should be managed equally, and early admission is required for avoiding severe pulmonary damage.

## Data Availability

NA

## Acknowledgements

The authors would like to show their respect for all the hospital staff for their hard work and efforts to combat the COVID-2019.

## Notes

### Competing Interest Statement

The authors have declared no competing interest.

### Funding Statement

No funding was provided for this study.

